# Whole genome characterisation of Enterotoxigenic *Escherichia coli* circulating in Zambia

**DOI:** 10.1101/2025.02.20.25322610

**Authors:** Suwilanji Silwamba, Michelo Simuyandi, Kapambwe Mwape, Charlie C Luchen, Kennedy Chibesa, Mwelwa Chibuye, Innocent Mwape, Fraser Liswaniso, Harriet Ngombe, Adriace Chauwa, Cynthia Mubanga, Sam Dougan, David Ojok, Andrew Moono, Nsofwa Sukwa, Monde Muyoyeta, Caroline C Chisenga, Roma Chilengi, Geoffrey Kwenda, Astrid von Mentzer

**Affiliations:** Centre for Infectious Disease Research in Zambia, Lusaka, Zambia; Department of Biomedical Sciences, School of Health Sciences, University of Zambia, Lusaka, Zambia; Water and Health Research Center, Faculty of Health Sciences, University of Johannesburg, P.O. Box 17011, Doornfontein, Johannesburg 2028, South Africa; Wellcome Sanger Institute, Hinxton, Cambridgeshire, CB10 1SA, UK; Department of Basic Medical Sciences, Michael Chilufya Sata School of Medicine, Copperbelt University, Ndola P.O. Box 71191, Zambia, Department of Microbiology and Immunology; Institute of Biomedicine, Sahlgrenska Academy, University of Gothenburg, Gothenburg, Sweden

**Keywords:** ETEC, Diarrheoa, Enterotoxins, Colonization Factors, Whole Genome Sequencing, Resistomes

## Abstract

**Background:** Enterotoxigenic *Escherichia coli* (ETEC) is a significant cause of diarrheal disease, particularly in low—and middle-income countries, including Zambia. ETEC pathogenesis is driven by colonisation factors (CF) and enterotoxins, and increasing antimicrobial resistance compounds the global health burden. Despite its impact, limited genomic data exists for ETEC strains in Sub-Saharan Africa.

**Methods:** This study conducted whole-genome sequencing of 62 ETEC isolates collected from children under five years old presenting with moderate-to-severe diarrhoea in Lusaka, Zambia. Genomic DNA was extracted, sequenced using the Illumina MiSeq platform, and analysed for phylogenetic relationships, toxin profiles, CF combinations, AMR genes, and plasmid incompatibility groups. Sequences were processed using bioinformatics tools, including SPAdes for genome assembly and Abricate for virulence and AMR profiling.

**Results:** The isolates displayed diverse phylogenetic groupings, predominantly within phylogroups A (22 isolates) and B1 (29 isolates). Forty-five serotypes and 39 sequence types were identified, with ST155, ST4, and ST847 being most prevalent. 35% of isolates lacked a known CF, but CS6-only stains were the most common (8%). The most frequent toxin profile was LTh (31%); AMR analysis revealed 350 resistance genes, with Sul2 (14%) and blaTEM-1 (10%) being predominant. ∼89% of isolates exhibited genomic multidrug resistance. Plasmid analysis identified IncFII as the most prevalent incompatibility group (19%).

**Conclusion:** This study highlights the genomic diversity of ETEC in Zambia, revealing concerns about multidrug resistance and identifying virulence profiles. These findings reiterate the urgent need for enhanced surveillance and targeted interventions, including vaccine development, to combat ETEC-related diarrhoea effectively.

**Authors Summary:** Diarrhoea caused by a bacteria called Escherichia coli is a significant health problem for young children in Zambia and many similar countries. Despite its profound impact, not much is known about the types of ETEC found in Zambia. In this study, we collected this type of bacteria from children under five years old who had diarrhoea and studied them in detail using advanced genetic tools. We discovered that the bacteria are very diverse, with different groups, toxin types, and ways of attaching to the host. Worryingly, most bacteria were resistant to several antibiotics, potentially making treatment challenging. This research helps us understand how ETEC spreads and causes illness in Zambia, showing the urgent need for tracking, improved treatments, and vaccines to protect children from this serious disease.

## Introduction

Enterotoxigenic *Escherichia coli* (ETEC) is endemic in low-middle-income countries (LMICs), where access to clean water and basic sanitation is a challenge [1]. The bacterium is one of the leading causes of moderate to severe diarrhoea and infects all age groups but commonly affects children and travellers to these endemic regions [2]. Globally, ETEC infections are estimated to cause 75 million diarrhoea episodes each year in children under 5 [3]. ETEC is responsible for morbidity and mortality rates of ∼ 4.2% of total diarrhoea-related deaths, thus remaining a public health concern [4]. Furthermore, long-term post-infection sequelae such as cognitive and developmental impairment have also been associated with repeated bouts of ETEC-associated diarrhoea [3,5].

*Escherichia coli* belongs to the family *Enterobacteriaceae* and forms six major clades: A, B1, B2, C, E, and F [6]. *E.coli* is a gram-negative rod-shaped bacillus with a genome size between 4.5 to 5.5 MB, and the genome has an average GC content of approximately 50% [7]. The bacterium is known to harbour plasmids that encode virulence and colonisation factors [8]. ETEC-mediated diarrhoea is contracted through the ingestion of contaminated food or water [9]. The pathogenesis of ETEC diarrhoea involves two main steps: (i) intestinal colonisation, followed by (ii) secretion of enterotoxins [10]. Either or both of two plasmid-encoded toxins, such as heat-stable enterotoxin heat-labile toxin (LT) or STa (STh and STp, encoded by the *estA* gene), which is genomically and structurally similar to the cholera toxin (CT) [11]. These toxins are secreted when the bacteria adhere to the target host epithelial cells, resulting in loss of ions and water characterised by the rapid onset of diarrheal illness [12].

ETEC colonisation factors (CFs) are fimbriae, fibrous structures or simple outer membrane proteins that facilitate adherence to the small intestinal mucosa [13]. Research has shown that the presence of certain CFs varies geographically [13]. ETEC cell interaction is host-specific, and with the CFs, the bacteria colonise the small intestinal epithelium outer membrane [13]. More than 25 different CFs have been identified and classified into four prominent families: CFA/I-like, CS5-like group, Class1b and a more diverse group of CFs [8,14]. Previous studies on the prevalence of different CFs have shown that at least 30% of the CFs from clinical ETEC isolates are yet to be identified[15] and are usually classified as CF-negative. Therefore, it is essential to identify the remaining novel CFs, as they play a vital role in the pathogenesis of the disease and are vaccine targets [15]. ETEC CFs are primarily encoded in the bacterial plasmid and are flanked by remnants of transposase or insertion sequences, suggesting they are acquired through horizontal gene transfer [16]. The common ETEC CFs and toxins combinations isolated from humans are CS7+LT, CS2 + CS3(±CS21) LT + STh, CS1 + CS3(±CS21) LT + STh and CS5 + CS6 LT + STh, CS6+ STp, CFA/I(±CS21) STh [15].

ETEC exhibits significant genetic diversity, with over 100 serotypes identified from clinical samples [17]. Whole Genome Sequencing (WGS) of a broad collection of global ETEC isolates has also identified 21 global lineages (L1–L21) [18,19]. Despite this extensive diversity, there is a notable correlation between lineage and colonisation factors such as enterotoxin production, CF, serotype, and plasmid content [20].

The escalating antimicrobial resistance (AMR) observed among ETEC and other enteric bacterial isolates poses a significant global challenge in combating these pathogens. The rapid emergence and dissemination of AMR-resistant bacteria outpace the development of replacement drugs, resulting in a reduction of the therapeutic arsenal. This, in turn, contributes to increased morbidity and mortality, further complicating the management of diarrhoeal diseases [21]. Furthermore, antimicrobial resistance patterns tend to vary regionally [21], yet there is a lack of data describing how these patterns have evolved, particularly in countries such as Zambia.

There is limited data on ETEC in Zambia as it has not been a pathogen of focus in enteric disease research and routine clinical screening. Simuyandi et al. and Silwamba et al. previously profiled CFs and enterotoxins in a Zambian study cohort using molecular techniques on retrospectively collected stool samples; however, they only profiled a select number of CFs. Colonisation factors are viable vaccine targets, and therefore, knowledge of the prevalence of colonisation factors using molecular or serological techniques could enhance the development of effective vaccines in regions with high ETEC diarrhoea [14,22]. This study aims to describe the genomic characterisation of prospectively isolated ETEC colonies from children presenting with moderate to severe diarrhoea at various health facilities in Lusaka, Zambia, profiling resistomes, putative virulence and colonisation factors.

## Methods

### Study Design

This was a laboratory-based cross-sectional study in which stool samples were collected from children with moderate to severe diarrhoea who sought care at health facilities in Lusaka, Zambia. Diarrhoea was specifically defined using the WHO standard as the occurrence of at least three loose or watery stools within 24 hours [23]. The obtained stool samples were submitted to the microbiology laboratory, where they were screened for the presence of ETEC.

### Study Sites

The study was conducted at five primary health facilities in peri-urban Lusaka: Matero Level 1 Hospital, George Clinic, Chawama Clinic, Kanyama Clinic, and Chainda South Clinic. These sites represent the peri-urban healthcare setting and serve diverse populations within their respective catchment areas.

Matero Level 1 Hospital, located in the Matero constituency, provides services to over 300,000 residents. George Clinic, positioned in the George compound, serves a catchment population of approximately 145,230 people. Chawama First Level Hospital, serving the Chawama compound, serves a population of about 200,000 residents. Kanyama First Level Hospital is in Kanyama township, a high-density, low-income residential area that serves nearly 400,000 inhabitants. Finally, Chainda South Clinic, situated near Chainda, kalikiliki and Mtendere Compounds, serves a smaller community of around 26,000 people within a settlement covering less than one square kilometre. Participants for the study were recruited from these health facilities as they accessed various health services provided at each location.

### Isolation and Identification of ETEC Strains

This study utilised archived and cryopreserved *E. coli* isolates stored at-80C [24]. After thawing, the isolates were sub-cultured on MacConkey agar (Oxoid, Hampshire, UK) and were then incubated for approximately 24 hours at 37°C. DNA extraction was performed on pure colonies through heat lysis, wherein a half-loopful of bacterial culture was emulsified in 100µl of molecular-grade water and boiled at 100°C for 10 minutes on a heating block. The resulting mixture underwent immediate centrifugation at 10,000 x g for 2 minutes, and 100µl of the supernatant was carefully added to a sterile 1.5 ml microfuge tube, which was then used as the DNA template for subsequent PCR reactions.

Multiplex PCR, targeting the LT, STh, and STp genes, was performed to confirm the identity of the ETEC strains using the specific primers shown in S1 Table. Briefly, the PCR reaction was conducted in a final volume of 20μl, comprised of 10μl of ReadyMixTM (x2) from the KAPA 2G Fast HS PCR kit (KAPA Biosystems, Merck, Darmstadt, Germany), 0.5μl of each *LT* (10mM) and *STp* (10mM) primer sets, 1μl of *STh* primer (10mM), 0.4μl of MgCl_2_ (25mM), 4.6μl of molecular-grade water, and 1μl of the DNA template obtained through rapid boil extraction. The ABI Gene Amp 9700 thermal cycler (AB Applied Biosystems, Foster City, CA, USA) was the instrument used for the PCR with the following cycling conditions: initial denaturation at 95°C for 2 minutes, then 30 cycles of denaturation at 95°C for 15 seconds, primer annealing at 52°C for 8 seconds, extension at 72°C for 10 seconds, and extension at 72°C for 2 minutes. Subsequently, all PCR amplicons were electrophoresed on a 1.5% agarose gel (Merck, Darmstadt, Germany) alongside a 100bp DNA Mass Ladder (ThermoFisher Scientific, Massachusetts, USA) for analysis[24].

### Genomic DNA Extraction and Quantification

ETEC isolate genomic DNA was extracted using the QIAamp DNA Mini Kit (QIAGEN, Hilden, Germany) following the manufacturer’s instructions and established protocols with a 60µl elution volume. DNA quantification was performed using the Qubit Fluorometer (ThermoFisher Scientific, Massachusetts, USA) to ensure accurate downstream analysis. Briefly, two assay tubes were set up for the standards, and one assay tube was used for each sample. The Qubit working solution (ThermoFisher Scientific, Massachusetts, USA) was prepared following the manufacturer’s instructions. Finally, the tubes were inserted into the Qubit Fluorometer for readings, with concentrations of 10ng/μl and above deemed adequate for NGS.

### DNA Library Preparation and Whole-Genome Sequencing

DNA libraries featuring an average insert size of 350bp were prepared using the Illumina DNA preparation kit (Illumina, San Diego, USA) and Miseq reagent kit v3 (Illumina, San Diego, USA) according to manufacturer’s instructions for subsequent sequencing on the Illumina MiSeq platform. The resultant reads average about 150bp in length, yielding approximately 1.2Gb of clean data for each isolate.

### Genome Assembly, Quality control and annotation

*De novo* assembly utilised the Shovill package and incorporated the SPAdes v3.14.1 algorithm (Bankevich et al., 2012). The assemblies’ sizes varied, ranging from 4.7 to 5.2 Mb, with an average size of 5.0 Mb, and exhibited a 50% GC content, which is deemed suitable for *E. coli*. Using CheckM2 [25], the evaluation indicated minimal contamination, with less than 0.5% of assemblies displaying any contamination beyond the predefined exclusions. For quality control purposes, the species identification of the sequenced isolates initially recognised as *E. coli* by Kraken/Bracken was subsequently advanced for further analysis. Assembled genomes were annotated by employing Bakta (https://github.com/oschwengers/bakta) [26] the Fastq sequences have been submitted to the NCBI database under Bioproject PRJNA1205103 and sequence accession numbers are shown in S2 Table.

## Statistical Analysis

### Baseline Characteristics

Summary statistics for all baseline variables were calculated. Where applicable, proportions and medians (IQR) were used to express categorical and continuous variables.

### Multi-locus Sequence Typing and serotyping

For the prediction of ETEC Multilocus Sequence Typing (MLST), the bioinformatic tool Abricate (https://github.com/tseemann/abricate) was employed, utilising the MLST tool available at (https://github.com/tseemann/mlst). Prediction parameters were established with a stringent threshold of 100% gene coverage and 60% identity. Similarly, for somatic O and flagella H antigen, an ETEC-specific database sourced from the Ecoh repository was used (https://github.com/katholt/srst2/tree/master/data).

### Identification of Enterotoxins, Colonisation Factors and Antimicrobial Resistance Genes

To predict the presence of virulence genes linked to ETEC, specifically to toxins (LTh, STh and STp) and colonisation factors, the bioinformatic tool Abricate was used (available at https://github.com/tseemann/abricate). The ETEC virulence and CF database employed in this study were sourced from Astrid von Mentzer’s repository, accessible at https://github.com/avonm/. Additionally, AMRfinderplus (https://github.com/ncbi/amr#ncbi-antimicrobial-resistance-gene-finder-amrfinderplus). A sample was considered positive for a specific CF if a complete set of genes, all corresponding to the same colonisation factor, were detected (major subunit, minor subunit, outer membrane usher and chaperone), an isolate lacking any of these genes from the database was considered to be CF negative. A 100% gene coverage threshold and 60% identity were applied to construct individual profiles for each sample. Simultaneously, toxin and resistome prediction was conducted using the same method on these samples. Schematic visualisation of percentage proportion analysis was done using R (ggplot 2). In the bubble charts generated by ‘geom_point’ in the ggplot2 package in RStudio, each circle represents a proportion, calculated as the number of genes divided by the total number of genes. The size of the circle corresponds to the relative abundance of the present gene.

### Plasmid incompatibility group analysis

To predict the presence of plasmid incompatibility groups found in the ETEC isolates, the frequency was determined using Plasmidfinder (https://cge.food.dtu.dk/services/PlasmidFinder/) and MOB-suite [27] bioinformatic tools were used to indicate the mobility of the plasmids.

### Phylogenetic Analysis

Roary (https://sanger-pathogens.github.io/Roary/) was employed to predict core genes across all samples. Following the identification of core genes, SNP differences among these genes were calculated using SNP-sites (https://github.com/sanger-pathogens/snp-sites).

Subsequently, a maximum likelihood tree was constructed utilising IQTREE (http://www.iqtree.org) with the GTR+F+I model, considering the computed fconst (123, 456, 789, 123). The visualisation of the generated tree was achieved using iTOL[28]

## Ethics Statement

The guardians or caregivers of the participants in this study gave consent on behalf of the children by filling in consent forms in commonly used languages in Lusaka. In cases of illiterate guardians, an impartial witness was used to interpret and provide a thumbprint signature as evidence of the agreement to the process as being voluntary. Participants’ recruitment period start and end dates were from 1^st^ September 2020 to 30^th^ November 2021. The study was conducted in line with ethical recommendations and requirements for the protection of human participants in research (HSP). The study obtained ethics approval from the University of Zambia Biomedical Research Ethics Committee (UNZABREC Ref: 1091-2020) and the National Health Research Authority (NHRA).

## Results

### Genomic characteristics of ETEC isolates collected in Zambia

A total of 62 confirmed ETEC isolates were sequenced from children under the age of 5 years presenting to the health facility with moderate to severe diarrhoea. The genomic characteristics of ETEC isolates collected in Zambia stratified by phylogroups. Isolates from Phylogroup A were collected from children with the highest mean age of one year and five months. The majority of the isolates belong to the phylogroup B1(n=29). Except for phylogroup C, all the other clades were detected in the Chainda location. Phylogroup B1 also had the most unique CF combination profiles (n=15), illustrated in Table 1.

**Table 1.** Genomic characteristics of ETEC isolates collected in Zambia.

| Phylogroup | No. of Isolates | Mean Age | Male (%) | Female (%) | Locations | Sequence Type | Toxin | Colonization Factor |
| --- | --- | --- | --- | --- | --- | --- | --- | --- |
| A | 22 | 1.48 | 40.91 | 31.82 | Chainda;<br>Kanyama;<br>George;<br>Matero;<br>Chawama | ST5348; ST1564; ST4;<br>ST4656; ST46; ST227; 0;<br>ST34; ST206; ST6438;<br>ST1491; ST226; ST165;<br>ST2332; ST6701; ST10;<br>ST1312 | LTh+STh; STh ; NO<br>TOXIN; LTh; STp | CS1+CS3+CS21; CF<br>Negative; CS2+CS3+ CS21;<br>CS6+CS21; CS31A;<br>CFA+CS21 |
| B1 | 29 | 1.21 | 48.28 | 41.38 | Chawama;<br>Matero;<br>George;<br>Kanyama;<br>Chainda | ST906; ST517; 0; ST443;<br>ST453; ST847; ST173;<br>ST1623; ST328; ST3857;<br>ST295; ST155; ST200; ST316;<br>ST9038 | LTh; STh; LTh+STp<br>; NO TOXIN; STp | CS6; CS14; CS13, CS26;<br>CS5+CS6; CS20+CS28A;<br>CS21; CS27B; CS7; CS27A;<br>CS17; CF Negative; CS13;<br>CS6+CS26+CS27A; CS28A |
| B2 | 2 | 0.9 | 50 | 50 | Chainda;<br>Chawama | ST452; ST28 | NO TOXIN | CF Negative |
| C | 2 | 1.05 | 100 | 0 | George | ST88 | LTh | CS7 |
| D | 2 | 2.6 | 100 | 0 | Chawama;<br>Chainda | ST69; 0 | NO TOXIN | CF Negative; CS23 |
| E | 2 | 1 | 0 | 100 | Chainda | ST3036; ST182 | NO TOXIN; STp | CF Negative; CS6 |
| F | 3 | 1.37 | 0 | 100 | George;<br>Chainda | ST3910; ST2141 | LTh+STp; NO<br>TOXIN | CS12; CF Negative |

Note: 27.27% and 9.74% of Gender information is missing for ETEC isolates belonging to phylogroup A and B1, respectively

### ETEC serotype and MLST frequency

Through whole-genome sequencing of the ETEC isolates, 45 unique serotypes based on the O and H antigens were identified. The most prevalent serotypes were O6:H16 4/62 (6%), O108var1:H27 3/62 (5%), O115:H5 3/62 (5%), and O92:H21 3/62 (5%) Fig 1A. This study identified 39 unique Sequence Types among the ETEC isolates derived from faecal samples. Novel ST, representing 5/62 (8%) of the isolates, were not previously catalogued in the Multi-Locus Sequence Typing (MLST) database. The most prevalent identified genotypes were ST155, ST4, and ST847 4/62(6.5%) Fig 1B

**Fig 1.**
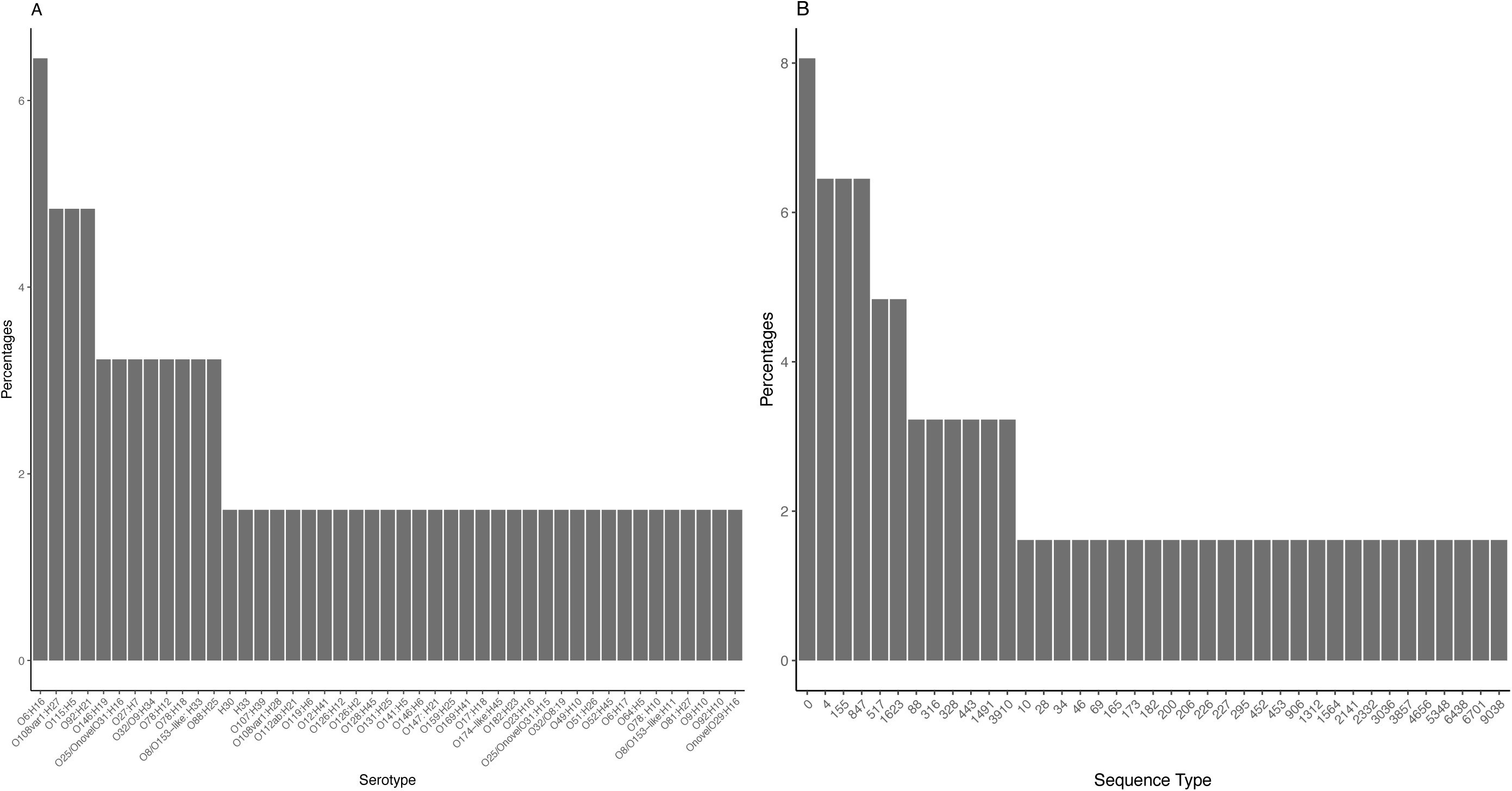
ETEC Serotype (A) and Sequence Type Frequency(B).

### Maximum Likelihood phylogenetic tree of global and Zambian ETEC strains

The global phylogenetic analysis of 424 ETEC genomes comprising 362 ETEC reference genomes reveals a significant diversity among both global and Zambian ETEC sequences, forming clusters across multiple lineages. Predominantly, the Zambian sequences cluster with phylogroups B1 and A, although there are notable representations in phylogroups B2, C, D, E, and F as well. A discernible trend emerges wherein strains sharing similarities in lineage, phylogroup, MLST, toxin profile, and colonisation factors tend to cluster together. The majority of CF negative strains cluster in new unidentified lineages, as shown in Fig 2

**Fig 2.**
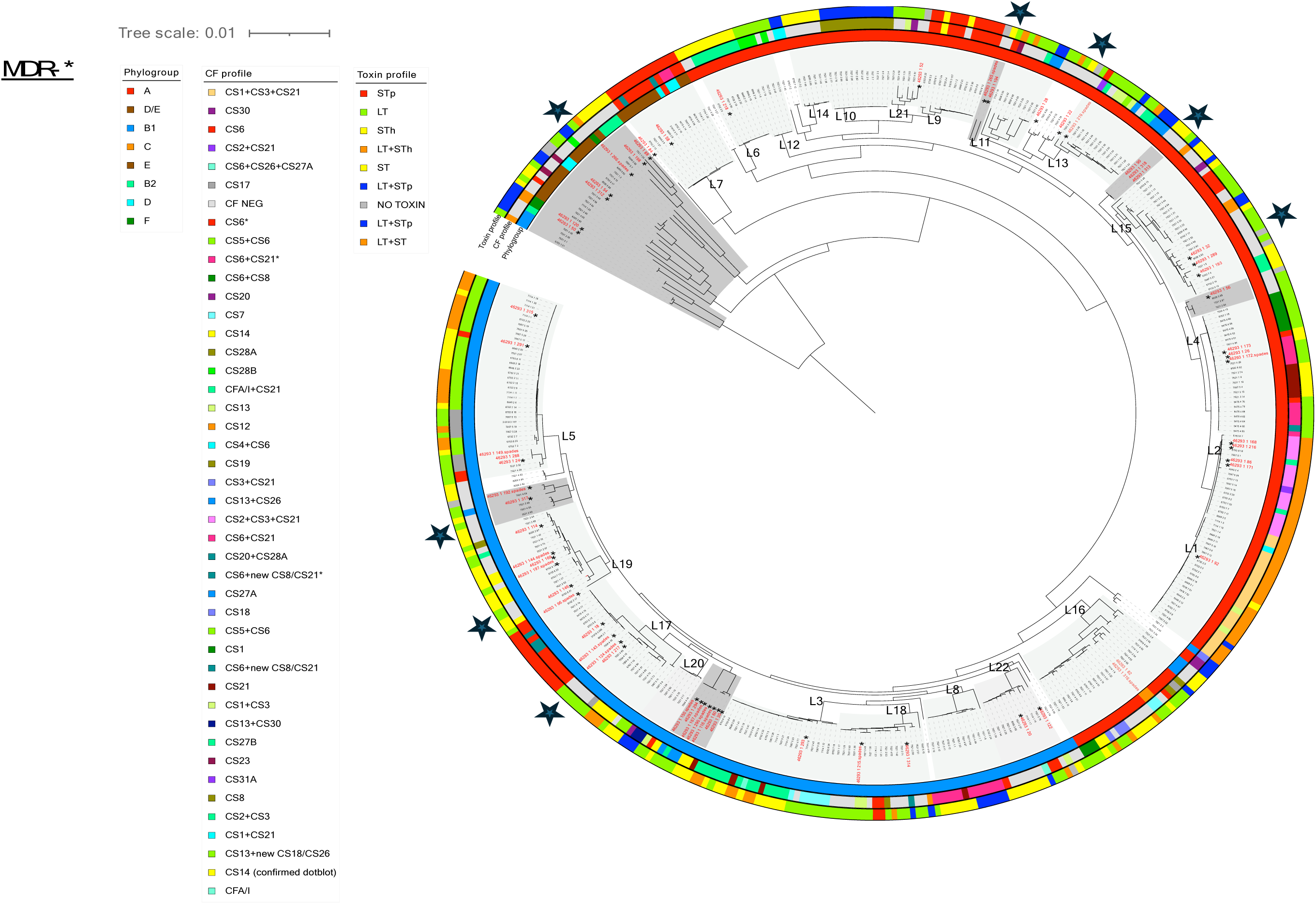
Maximum Likelihood phylogenetic tree of global ETEC strains. Note: * represents Multidrug resistance (MDR) Zambian ETEC isolates, and STARS show CF-negative stains location; dark grey shades represent new/unidentified lineages

### ETEC WGS toxin gene distribution

Through whole genome sequencing analysis of the 62 ETEC isolates. The most prevalent toxin profile was LTh 19/62 (31%). The second most frequent toxin gene observed was STh 14/62 (22%), followed by combinations of LTh+ STh 5/62 (8%), LTh+STp 4/62 (6.5%), and STp only 4/62 (6.5%) Fig 3A. Although 15 isolates were PCR toxin positive, however negative on WGS analysis Fig 3B

**Fig 3.**
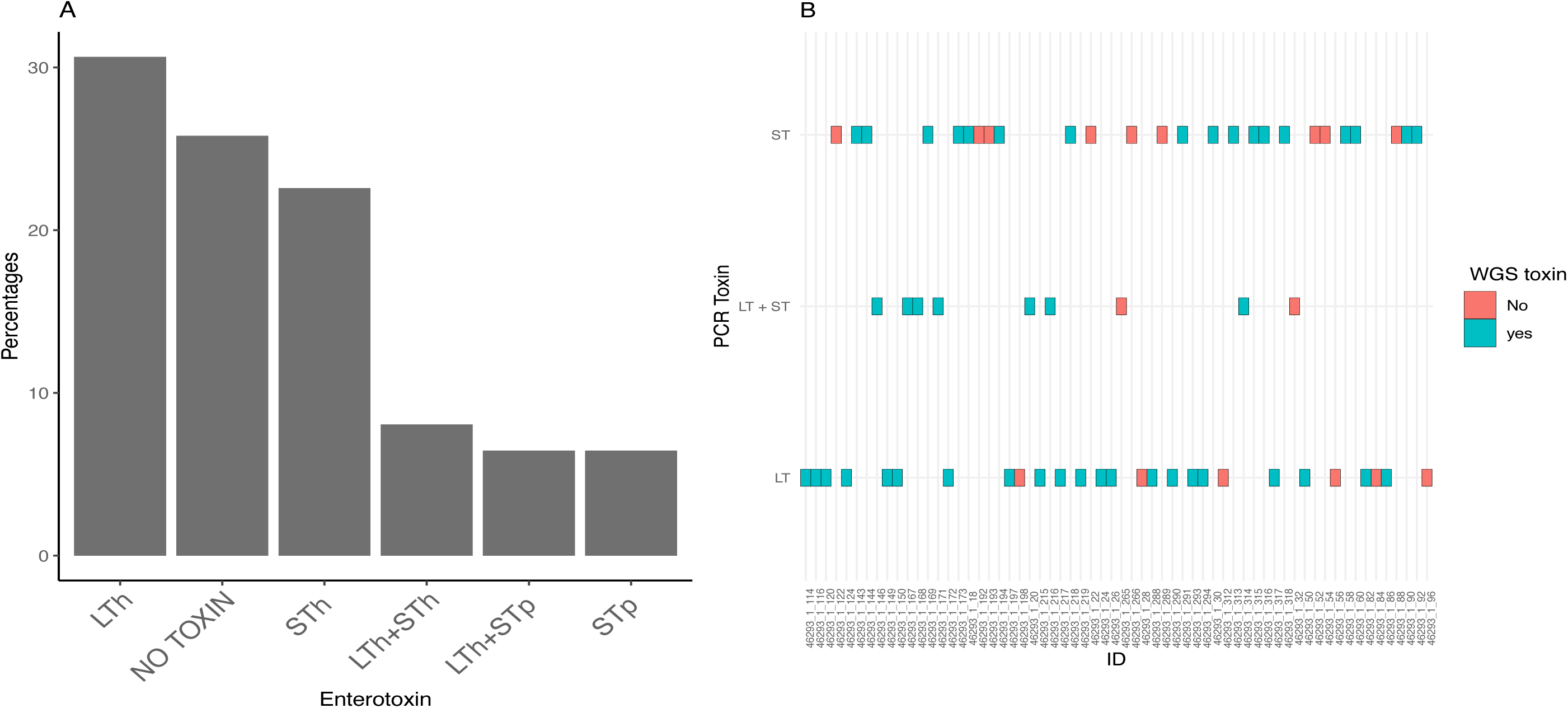
WGS enterotoxin gene distribution (A). Laboratory PCR and WGS enterotoxin detection comparison (B)

### Colonisation factor frequency

Zambian ETEC whole-genome sequencing identified 20 strain combinations of colonisation factors (CFs), excluding those unidentified or absent in current databases from the 62 isolates. The majority of the sequenced ETEC isolates lack a known CF 22/62 (35%). Among the identified CFs, CS6-only strains were the most prevalent, 5/62 (8%). Other combinations of CFs identified included CS14 4/62 (6%), CS2+CS3+CS21 4/62 (6%), CS17 3/62 (5%), CS27A-only 3/62 (5%), CS6+CS21 3/62 (5%), CS7 3/62 (5%) Fig 4A. post-stratification of each unique CF, 59 different colonisation factors were identified, excluding 22 that were unidentified or not present in current databases. Among the identified CFs, CS6 and CS21 were the most prevalent 10/81 (12.3%), respectively Fig 4B. The frequency of Fimbrial Usher Protein (FUP) clades of each unique CF was calculated, and three different individual FUPs were identified based on [8] Among the identified FUPs, Gamma were the most prevalent 24/77 (31.2%). Other FUPs identified included Alpha 17/77 (22.1%), Longus 9/77 (11.6%) and 5/77(6.5%), which fall into the diverse group of kappa FUPs shown in Fig 4C.

**Fig 4.**
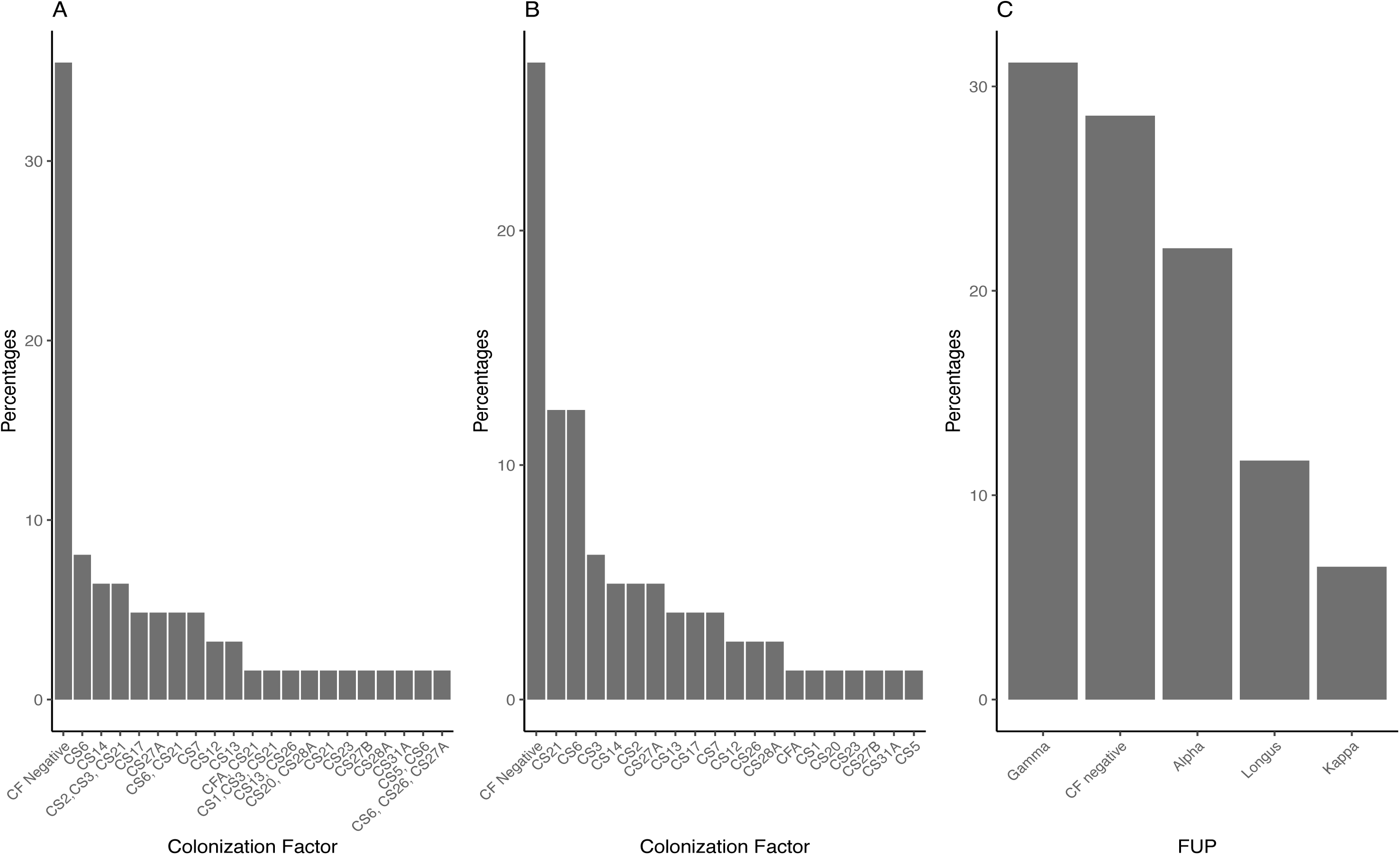
Colonisation factor strain profile frequency (A) ETEC colonisation factor cumulative frequency (B). Fimbrial Usher Protein (FUP) frequency (C)

### Antimicrobial resistomes

A total of 350 antimicrobial resistance genes (ARGs) were identified from the whole-genome sequences of ETEC. The most prevalent resistance gene detected was *Sul2*, which accounted for 49/350 (14%) of the total genes. *Sul2* encodes a Transposase that confers resistance to Trimethoprim/Sulfamethoxazole. Other ARGs included *blaTEM-1* 36/350 (10%), *aph(6)-Id* 33/350 (9%), *blaEC-18* 26/350 (7%), *aph(3’’)-Ib* 22/350 (6%), *dfrA14* 19/350 (5%), *aadA1* 16/350 (5%), *tet(A)* 16/350 (5%), *blaEC-15* 15/350 (4%), *sul1* 14/350 (4%), *qnrS1* 13/350 (4%), *dfrA1* 11/350 (3%), *dfrA8* 11/350 (3%), *blaEC* 8/350 (2*%), tet(B)* 8/350 (2%), *blaEC-13* 7/350 (2%), *dfrA7* 6/350 (2%) (Fig 5A). All isolates (100%) exhibited resistance to cephalosporins. In total, 11 antibiotic drug classes were detected, and 88.7% (55/62) of the isolates had genes that encoded resistance to at least three antibiotic classes (Fig 5B).

**Fig 5.**
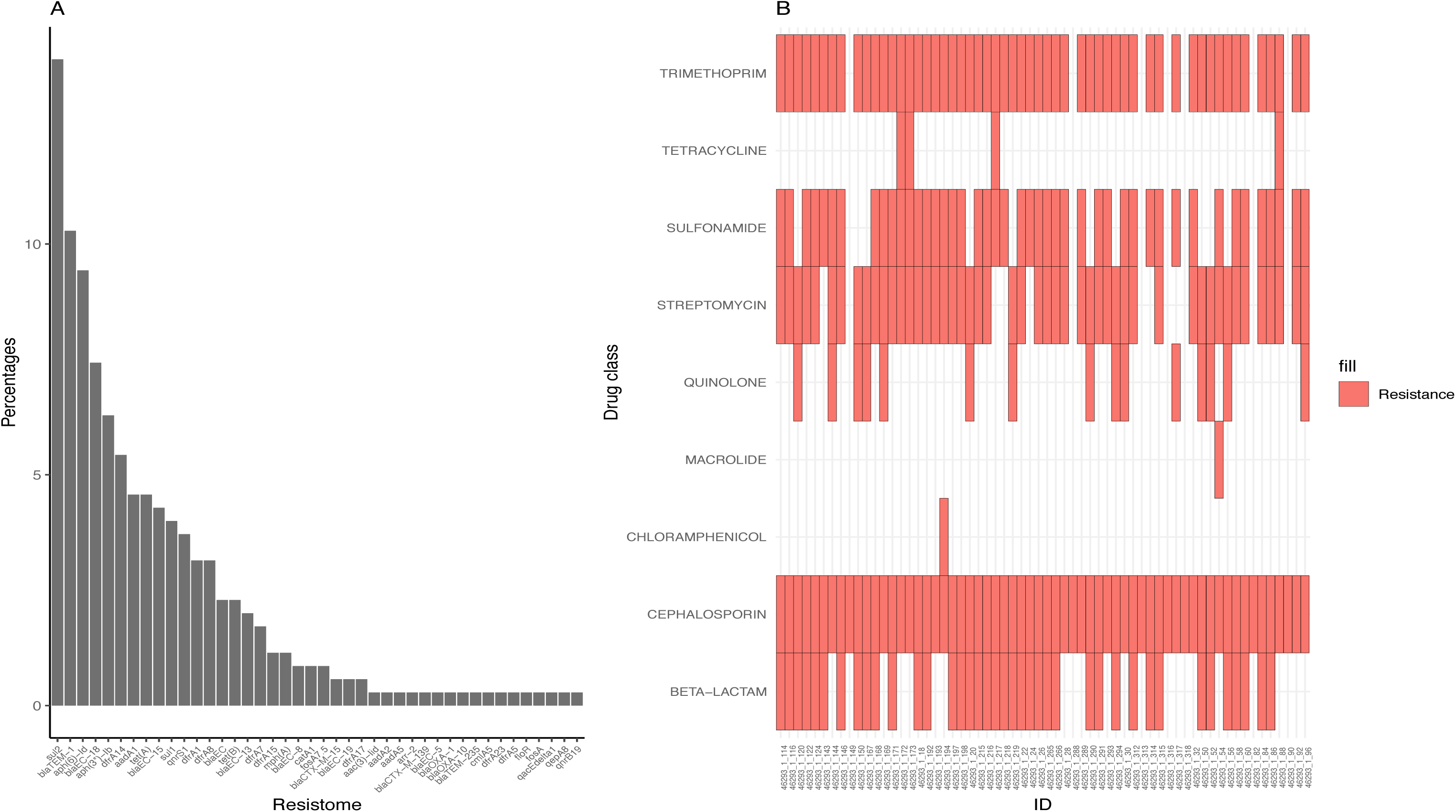
Antimicrobial resistome frequency(A) Detection of multi-drug resistance strains (B)

### Interplay Between Colonization Factors and Antimicrobial Resistance Drug Classes

It was observed that every ETEC isolate (100%) exhibited resistance to cephalosporin strains possessing CS7, CS21, CS13+CS26, and CS1+CS3+21 colonisation factors showed no resistance to beta-lactams, with cephalosporin resistance being the most prevalent among the resistant strains. Fosfomycin resistance was exclusively identified in strains with CS27A and CS27B colonisation factors, while quinolone resistance was predominantly associated with gamma variants. Interestingly, CF-negative strains displayed resistance to all antimicrobial drug classes except fosfomycin. Specifically, CS1 and CS7 strains lacked beta-lactam antimicrobial resistance genes. Similarly, CS20 and CS17 strains did not exhibit streptomycin resistance genes, as shown in Fig 6.

**Fig 6.**
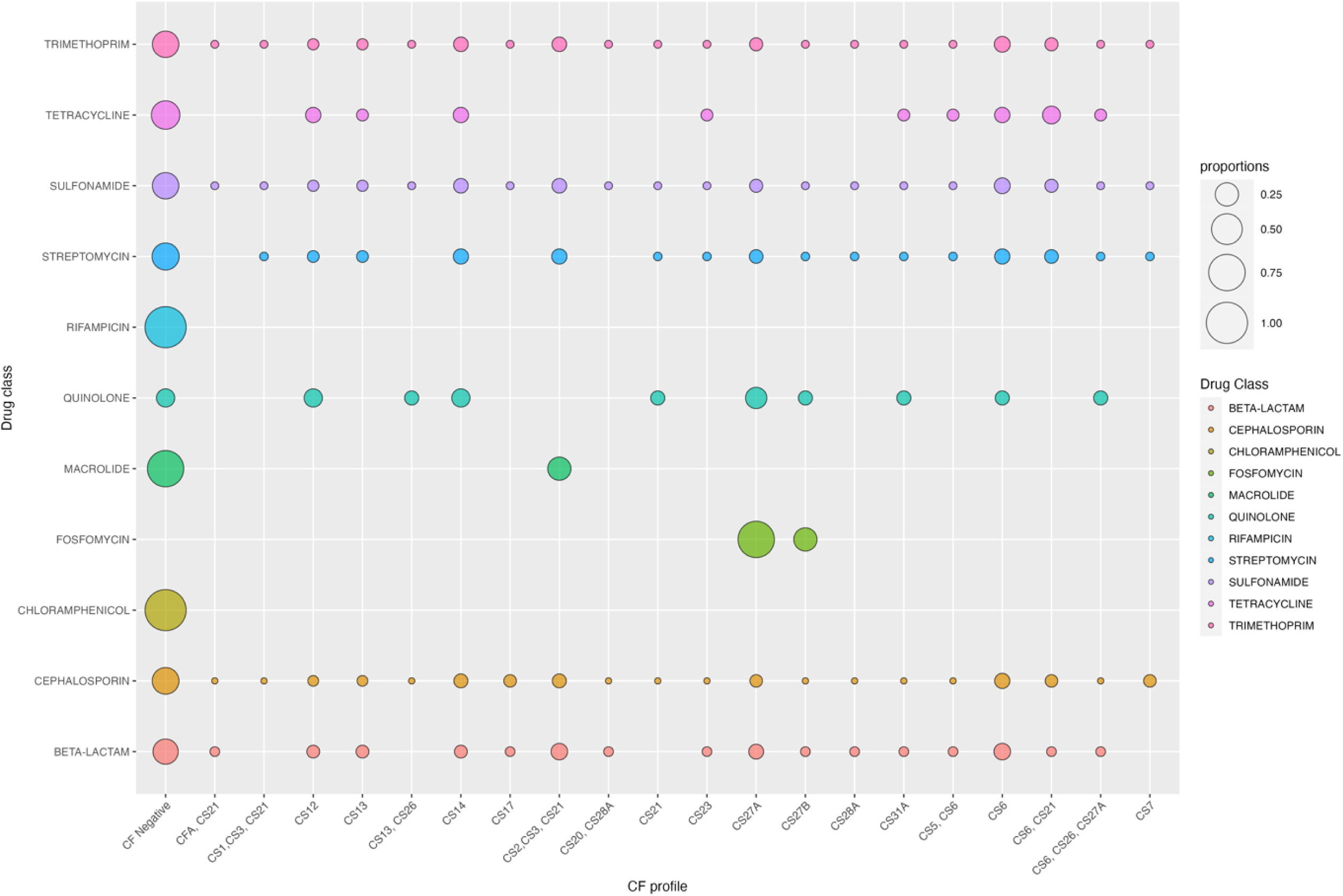
Association between Colonization Factors and Antimicrobial Resistance Drug class

### Plasmid Incompatibility Group Frequency

The whole-genome sequences of the 62 ETEC isolates identified 223 plasmid incompatibility groups. The most prevalent incompatibility group detected was IncFII, which accounted for 42/223 (19%) of the total plasmids (S1 Fig), and the majority of plasmids identified are conjugative plasmids harbouring multiple resistomes. All the Rep_cluster 2335 plasmids are predicted mobilisable and hobour resistomes for aminoglycosides, trimethoprim and quinolones. *blaCTX-M-139, blaCTX-M-15* and *drfA5* exclusively found in incI-gamma/k1 shown in Fig 7.

**Fig 7.**
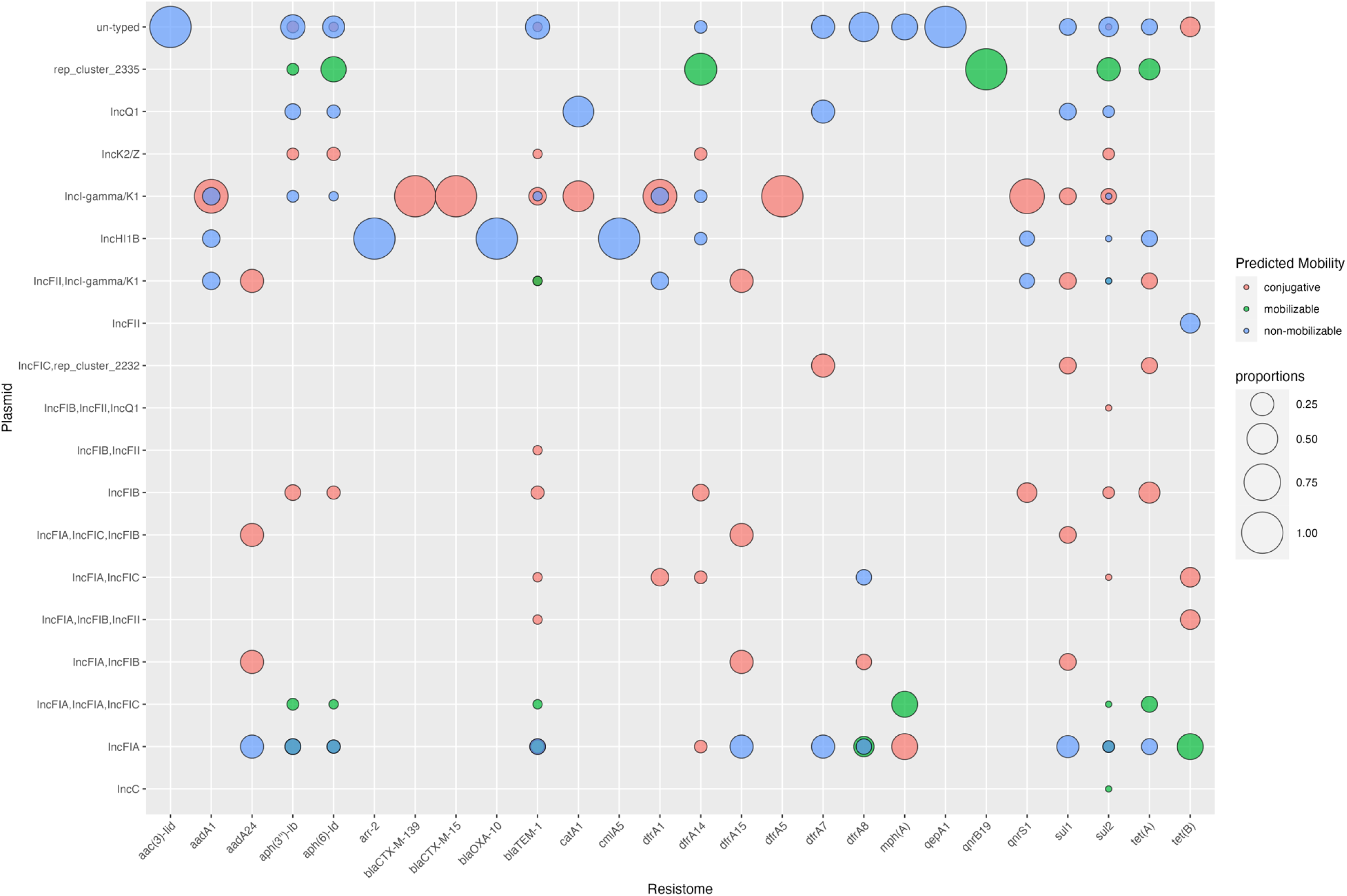
Association between Plasmid Incompatibility Group and Resistomes

## Discussion

This study reconstructs the genomic population structure of ETEC Zambian isolates using WGS, placing them within the broader framework of global ETEC lineages. Additionally, it characterised the virulence factors and AMR mutation/gene profiles of the Zambian isolates. To our knowledge, this represents the first investigation into the genomic epidemiology and AMR profile of ETEC infections in Zambia.

The findings revealed a concerning prevalence of antibiotic resistomes, with the majority of isolates demonstrating resistance to more than one antibiotic class. Notably, most isolates exhibited at least one resistome associated with cephalosporin, trimethoprim, and sulfonamide resistance, indicating widespread resistance within the sampled population. ∼88% of the isolates are potentially multi drug resistance harbouring ARGs that could cause resistance to at least three antibiotic classes. These findings are similar to reports from a previous study evaluating AMR of *E.coli* obtained from food, animal, and human sources, which shows a concern for cephalosporin resistance [29].

Additionally, substantial resistance was detected against streptomycin, beta-lactam, tetracycline, and quinolones, further complicating treatment options. These findings are similar to those previously described by Chiyangi et al., profiling 15 DEC isolates in Zambia exhibiting significant resistance to cotrimoxazole, ampicillin, cefotaxime, ceftazidime, cefpodoxime, nalidixic acid, and tetracycline [30]. The most prevalent resistance gene observed was Sul2. Sul2 encodes a transposase that confers resistance to Trimethoprim/Sulfamethoxazole, a commonly used antibiotic [31]. The high prevalence of Sul2 could be attributed to the usage of Septrin (cotrimoxazole) in an HIV-endemic population as a prophylaxis for HIV-related infections among adults, adolescents and children.

Several other ARGs were identified, including *blaTEM-1, blaEC-18*, and *blaEC-15*. The study demonstrated that the prevalence of Extended-Spectrum Beta-Lactamase (ESBL)-producing ETEC isolates among outpatients in Lusaka (32%) was comparable to post-2013 rates among diarrhoeal patients in Nepal (30%) [32] and among travel cases to Southeast Asia/India (43%) [33]. These findings could inform treatment management of travellers to ETEC endemic areas in sub-Saharan Africa.

The analysis of plasmid incompatibility groups revealed an array of plasmid types indicative of horizontal gene transfer and the potential distribution of antibiotic resistance and virulence genes. Among the plasmid incompatibility groups identified, IncFII emerged as the most prevalent. This finding is significant as IncFII plasmids are known to carry various antibiotic resistance genes and virulence factors, contributing to the adaptability and pathogenicity of ETEC strains [34]. These findings highlight the repertoire of plasmids harboured by ETEC isolates, each potentially playing a role in mediating antibiotic resistance or other survival adaptive traits [35]. Understanding the distribution and prevalence of plasmid incompatibility groups is crucial for elucidating the mechanisms involved in the acquisition of antibiotic resistance and virulence determinants among ETEC populations.

The findings from this study shed light on the intricate relationship between colonisation factors and antimicrobial resistance patterns in ETEC isolates. An analysis revealed associations between specific colonisation factors and resistance profiles. Intriguingly, strains harbouring certain colonisation factors exhibited distinct resistance patterns. For instance, strains carrying CS7, CS21, CS13+CS26, and CS1+CS3+21 colonisation factors demonstrated susceptibility to beta-lactams, suggesting a potential correlation between these CFs and antimicrobial susceptibility similar to a paper published in Nepal [32]. Conversely, resistomes to fosfomycin were exclusively identified in strains possessing CS27A and CS27B colonisation factors, indicating a potential association between these factors and fosfomycin resistance.

Furthermore, The study identified other distinct resistomes exclusive to certain CF variants (S4 Fig). For instance, the qnrB19 and *blaTEM-235* resistomes were exclusively identified in CS31A ETEC strains, and the *blaCTX-M-139* resistome was found in the CS6+CS26+CS27A ETEC strain, emphasising the diversity of resistance mechanisms among different CF combinations. The absence of carbapenemase resistance genes across all examined isolates is a reassuring finding, suggesting that carbapenems remain effective against ETEC strains.

Several unique serotypes were identified based on the somatic O and flagella H antigens, further highlighting the genetic diversity among ETEC strains. Among the identified serotypes, some emerged as the most prevalent in this study cohort. The most frequently encountered serotypes were O6:H16, O108var1:H27, O115:H5, and O92:H21. The ETEC serotype O6:H16 has been implicated in several outbreaks in Japan over the years, including a large outbreak (510 cases in 2005) linked to the consumption of vegetables [36]. These findings emphasise the importance of these serotypes as significant contributors to the burden of ETEC infections. Interestingly, in the past, ETEC serotypes of O27:H7 were isolated from diarrheic patients in Brazil [37]. While less prevalent than the dominant serotypes, these findings highlight the wide range of serotypic diversity in the ETEC population, as shown in the results.

In this study, we identified a diverse array of sequence types; however, most identified sequence types were novel, with the isolates exhibiting uncatalogued STs in the MLST database. ST10 was also observed to be the common ancestor of all the STs (S2 Fig), similar to previously reported [19]. This highlights the importance of ongoing surveillance efforts to capture emerging genetic variants of ETEC evolution. Among the identified sequence types, several emerged as the most prevalent in this study. ST155, ST4, and ST847 were the most frequently encountered genotypes of the isolates. The Zambian strains ST4 exhibited the same toxin and CF profiles as those isolated from Bolivia’s clinical samples within the same phylogroup [38]. These findings suggest the potential dominance of these sequence types within specific geographic regions or populations and may have implications for understanding the epidemiology of ETEC infections. ETEC O169:H41 (ST182) strains are acknowledged for their association with traveller’s diarrhoea and foodborne outbreaks in developed countries, prompting considerable concern [39]. A recent study obtained draft genomes of O169:H41 (ST182) E. coli from two aeroplane waste samples gathered at a German international airport [39]. It is noteworthy that all ST182 isolates were classified into phylogroup E, reflecting the phylogenetic classification of the ETEC isolate discovered in Zambia, and displayed identical toxin and colonisation factor profiles.

The study revealed that the Zambian sequences are dispersed among seven distinct phylogroups, with phylogroups A and B1 emerging as the most predominant. Furthermore, the Zambian ETEC sequences also exhibit affiliation to other representations in phylogroups B2, C, D, E, and F. These findings are similar to several studies reporting the high prevalence of these ETEC phylogroups >40% of B1 in Bangladesh [40] and a high prevalence of phylogroup A in Chile and China [41,42].

Furthermore, identifying multiple phylogroups among the Zambian sequences suggests a diverse array of ETEC strains circulating within the population. Zambian isolates with serotype O6:H16 and MLST-ST 4 exhibited a virulence factor profile similar to that of L2 in the global dataset, characterised by the presence of STh + LT and CS2 + CS3, with the possible presence of CS21. The serotype and MLST-ST of the L2/4 lineage were determined as O6:H16 and ST48, respectively. ETEC isolates with this particular serotype and MLST-ST combination have been previously reported in pigs in Denmark [43] and human cases in China [44]. Of particular interest is the observation that strains sharing similar toxin, MLST, CF, and AMR profiles tend to cluster together phylogenetically [42,45]. A noteworthy finding is that despite identifying ETEC strains via PCR, a few isolates lacked associated toxin genes. A previous research publication observed a similar occurrence of losing virulence plasmid of an unstable *E.coli* coding for STp+CS6 after an overnight passage [46]. Remarkably, these sequences still form clades with global reference ETEC genomes, indicating that other genetic factors beyond traditional toxin genes may contribute to their evolutionary history.

Of particular interest is identifying distinct profiles, such as CS2+CS3+CS21+LT+STh, which collectively form a distinct profile known as L1 seen globally and could indicate an “outbreak” of commonly spread ETEC [18]. A particularly intriguing observation is the identification of seven Zambian genomes forming two unique lineages stemming from lineage L20, sharing similar toxin profiles (*LT*) and colonisation factors (CS27A and CS27B). This suggests a potential clonal expansion or localised transmission of specific ETEC strains within the Zambian population. Finally, the phylogenetic analysis indicates that strains belonging to phylogroup B1 are distantly related to strains belonging to phylogroups B2, D, E, and F.

One striking observation from the analysis is that most of the sequenced ETEC isolates lack a known CF. These findings support reports from a review paper that stated that 30% of clinical ETEC isolates lack a known colonisation factor [47]. This suggests the presence of novel or unidentified CFs within the Zambian ETEC population. Utilising the updated classification for colonisation factors (CFs) proposed by Von Mentzer et al., [8] which is based on protein sequence comparisons of conserved outer-membrane usher proteins, this study classified CFs into fimbrial usher protein (FUP) clusters. The findings revealed that ETEC Gamma variants are the predominant strains circulating within this population.

Among the identified CFs, strains possessing CS6-only were the most prevalent. CS6 is a well-known colonisation factor associated with ETEC pathogenesis, facilitating adherence to host intestinal epithelial cells [46]. Its high prevalence in this study is vital in managing ETEC infections in Zambia, especially as a component in the ETVAX vaccine formulation. In contrast to the findings of The Global Enteric Multicenter Study, which reported a high prevalence of CFA/I-ETEC strains, this specific colonisation factor profile was minimal within the Zambian population [48]. These highlight the geographical variability of colonisation factors.

The study found that the *LTh* gene was the most prevalent enterotoxin, followed by *STh* and *LTh+STh*. Other toxin combinations were the least prevalent, such as *LTh+STp* and *STp* only. These findings are similar to those observed in a pilot study in Zambia [22,49].

In descending order of frequency, the distribution of toxin and CF combinations revealed several notable patterns (S3 Fig). The most prevalent combination was STh+CF negative, followed by LTh+STh+CS2+CS3+CS21, *STp*+CS6 and STh+CS14. Interestingly, strains with only the *LTh* gene were found to possess a broader spectrum of known colonisation factors compared to those with only *STh* genes. The toxin profiles of ETEC are typically associated with specific colonisation factors. For instance, combinations such as CFA/I+ST, LT+ST+CS1+CS3, or LT+ST+CS2+CS3 are often observed [14]. These findings provide valuable insights into the epidemiology and virulence characteristics of ETEC strains circulating within the studied population. Additionally, we identified strains presenting with toxin and CF combinations that have yet to be extensively documented in previous studies, highlighting the genetic diversity and potential for novel pathogenic profiles among ETEC isolates. Although 30%–40% of all ETEC strains remained negative for any CF, research on CF-negative strains has led to the discovery of new CFs, such as CS23, and an opportunity for further research studies[38,50].

## Limitations

This study was limited to ETEC isolates collected from outpatient health facilities, which may have excluded more severe cases typically found in inpatient settings. The small sample size could have limited the capture of the full genomic diversity of ETEC. Although ETEC was genetically characterised, phenotypic AMR expression was not evaluated; only the presence of ARGs was assessed. Consequently, not all genes detected are expressed phenotypically, although external pressures could activate those not currently expressed.

## Conclusion

In conclusion, by employing whole genome sequencing alongside traditional molecular subtyping methods, valuable insights were gained into the intricate relationships between ETEC isolates, their toxin profiles, colonisation factors, and antimicrobial resistance mechanisms. Firstly, the study highlights the limited understanding of ETEC infections in children, emphasising the need for advanced molecular techniques like WGS to understand ETEC epidemiology. The genomic analysis revealed a concerning prevalence of antimicrobial resistance genes among ETEC isolates, with widespread resistance observed across multiple antibiotic classes. This highlights the urgent need for surveillance and stewardship efforts to mitigate the emergence of antimicrobial resistance in ETEC populations. Moreover, the study identified distinct phylogenetic lineages, serotypes, sequence types and plasmid incompatibility groups among Zambian ETEC isolates, describing the genetic diversity within the population. Continued research efforts are essential to stay ahead of evolving ETEC strains and effectively mitigate the burden of diarrhoeal disease worldwide.

## Funding

Astrid von Mentzer was supported by The Swedish Research Council (grant no. 2022-01449) and the Sahlgrenska Academy International Starting Grant. This project is part of the European and Developing Countries Clinical Trials Partnership (EDCTP) program, supported by the European Union (grant number RIA 2018V-2309—ETEC, ETVAX).

## Author contributions

The authors contributed significantly to various aspects of this study, reflecting a collaborative effort. SS, MS, RC, GK, and AVM were instrumental in conceptualising the study. SS, CCL, SD, KC, and AVM conducted the formal analysis, ensuring the accuracy of the data interpretation. SS, KM, KC, CM, FL, AM, IM, SD led methodology. SS, MS, CCC, FL, HN, AC, CM, KM, IM, AM, and NS led the writing of the original draft. SS, KM, DO, AM, IM, FL, NS, MC, and KC spearheaded investigation efforts, including data collection and laboratory analysis. CCC, MS, MM, GK, and AVM provided supervision during the research. Finally, SS, MS, KM, CCL, KC, MC, IM, FL, HN, AC, CM, SD, DO, AM, NS, MM, CCC, RC, GK, and AVM collectively reviewed and edited the manuscript.

## Data Availability Statement

Data cannot be shared publicly to protect the confidentiality of study participants. However, researchers can inquire on accessing the metadata by requesting from the Centre for Infectious Disease Research in Zambia (CIDRZ) Institutional Data Access/Ethics Committee by contacting. All sequence data has been submitted to the NCBI database.

## Acknowledgements

We gratefully acknowledge the European and Developing Countries Clinical Trials Partnership (EDCTP) for their financial support, which made this project possible. Our sincere thanks also go to the Wellcome Sanger Institute for their invaluable technical assistance. Additionally, we extend our gratitude to the University of Gothenburg and the dedicated CIDRZ team for their unwavering commitment and contributions to the successful completion of this work.

## Notes

### Competing Interest Statement

The authors have declared no competing interest.

### Funding Statement

AVM was supported by The Swedish Research Council (grant no. 2022-01449) and the Sahlgrenska Academy International Starting Grant. RC was awarded funding for this project as part of the European and Developing Countries Clinical Trials Partnership (EDCTP) program, supported by the European Union (grant number RIA 2018V-2309—ETEC, ETVAX). The funders had no role in study design, data collection and analysis, decision to publish, or preparation of the manuscript

### Author Declarations

The study was conducted in line with ethical recommendations and requirements for the protection of human participants in research (HSP). The study obtained ethics approval from the University of Zambia Biomedical Research Ethics Committee (UNZABREC Ref: 1091-2020) and the National Health Research Authority (NHRA).

